# Contribution of infection and vaccination to seroprevalence through two COVID waves in Tamil Nadu, India

**DOI:** 10.1101/2021.11.14.21265758

**Authors:** T.S. Selvavinayagam, A. Somasundaram, Jerard Maria Selvam, Sabareesh Ramachandran, P. Sampath, V. Vijayalakshmi, C. Ajith Brabhu Kumar, Sudharshini Subramaniam, K. Parthipan, S. Raju, R. Avudaiselvi, V. Prakash, N. Yogananth, Gurunathan Subramanian, A. Roshini, D.N. Dhiliban, Sofia Imad, Vaidehi Tandel, Rajeswari Parasa, Stuti Sachdeva, Anup Malani

## Abstract

Four rounds of serological surveys were conducted, spanning two COVID waves (October 2020 and April-May 2021), in Tamil Nadu (population 72 million) state in India. Each round included representative populations in each district of the state, totaling ≥20,000 persons per round. State-level seroprevalence was 31.5% in round 1 (October-November 2020), after India’s first COVID wave. Seroprevalence fell to 22.9% in 2 (April 2021), consistent with waning of antibodies from natural infection. Seroprevalence rose to 67.1% by round 3 (June-July 2021), reflecting infections from the Delta-variant induced second COVID wave. Seroprevalence rose to 93.1% by round 4 (December 2021-January 2022), reflecting higher vaccination rates. Antibodies also appear to wane after vaccination. Seroprevalence in urban areas was higher than in rural areas, but the gap shrunk over time (35.7 v. 25.7% in round 1, 89.8% v. 91.4% in round 4) as the epidemic spread even in low-density rural areas.

**Article Summary Line:** Antibodies waned after India’s first COVID wave and both vaccination and infection contributed its roughly 90% seroprevalence after its second wave.

## Introduction

Knowledge of population-level immunity is critical for understanding the epidemiology of SARS-CoV-2 (COVID-19) and formulating effective infection control, including the allocation of scarce vaccines. Tamil Nadu is the 6th most populous state in India, with roughly 72 million persons *(1)*. India, including Tamil Nadu, experienced three COVID-19 waves that peaked in September 2020, May 2021, and February 2022 *(2)*. India has reported 43 million COVID-19 cases and 524,000 COVID-19 deaths through May 31, 2022 *(2)*. Tamil Nadu has reported roughly 3.4 million COVID-19 cases and 38,000 deaths, ranked 4th highest among Indian states through May 31, 2022 *(3)*. Reported cases are not, however, gathered from population-representative samples. Moreover, low testing rates may cause cases to underestimate population-level immunity.

To address these concerns, the state government conducted population-level serological surveys in 4 rounds, in October-November 2020, April 2021, June-July 2021, December 2021-January 2022 (Figure 1). Each survey was conducted on representative populations in each district of the state, except Chennai in round 2. We report seroprevalence estimates from these surveys by district, by demographic groups, and by urban status. We compare the results of the surveys to estimates from reported cases to measure the degree to which reported cases underestimate population immunity. We examine the extent to which infection and vaccination contributes to seroprevalence by comparing rates of infection and vaccination to changes in seroprevalence across rounds of surveys. We infer the extent to which antibodies decline following infection and vaccination by using data on changes in district-level seroprevalence across rounds and individual reports of the date of their own infection and vaccination, respectively.

**Figure 1.**
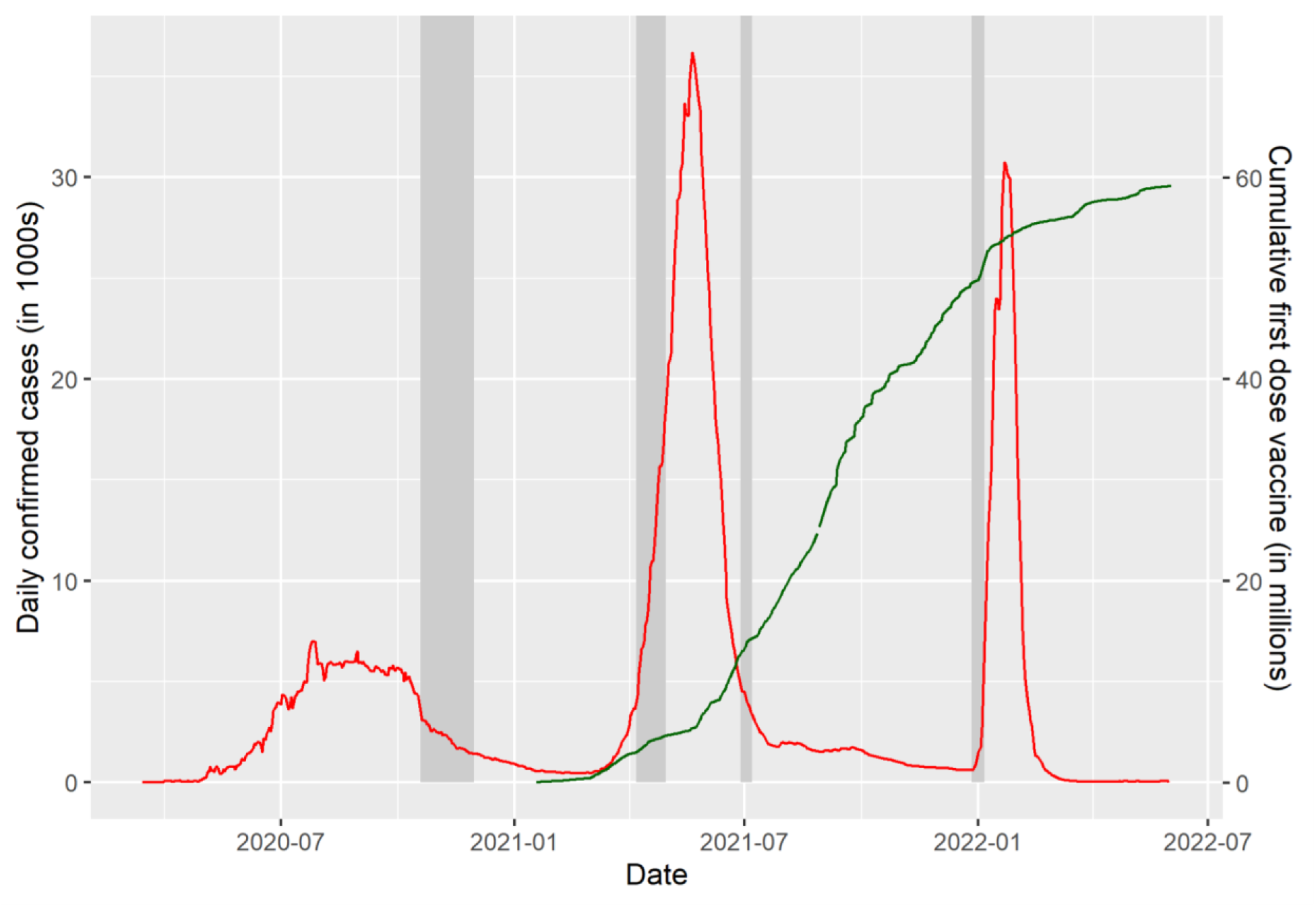
Daily new infections, new vaccinations (dose 1), and dates of serological survey rounds in Tamil Nadu

## Methods

The study was approved by the Directorate of Public Health and Preventive Medicine, Government of Tamil Nadu, and the Institutional Ethics Committee of Madras Medical College, Chennai, India. The study was entirely funded by the Government of Tamil Nadu and the National Health Mission, Tamil Nadu.

### Outcomes

The primary endpoints are (1) the fraction of the population that would obtain positive results on CLIA (chemiluminescent immunoassay) antibody tests for COVID, i.e., seropositivity, at the district-level, and (2) the fraction of the population that have antibodies for COVID, i.e., seroprevalence, district level.

The secondary endpoints are (1) seroprevalence (a) by age and sex, (b) by urban status, and (c) at the state level; (2) the difference between population immunity estimated by serological survey and by reported cases; and (3) self-reported infection and vaccination.

### Survey timing, sample, and location

Data was gathered between 19 October – 30 November 2020, 7 – 30 April 2021, 28 June to 7 July 2021, 27 December 2021 – 6 January 2022 in rounds 1, 2, 3 and 4, respectively. Individuals residing in Tamil Nadu and ages 18 years and older were eligible for rounds 1 to 3 of this study. In round 4, eligibility was expanded to ages 10 and older. The exclusion criteria were refusal to consent and contraindication to venipuncture. In round 2, Chennai district was not surveyed because there was an outbreak that prevented sampling in that district.

### Sample size

Sample sizes for rounds 1 to 3 were calculated assuming a seropositivity of 0.5 throughout the state, to maximize sample size. For round 4, the positivity rate estimated from round 3 (0.662) was used. Calculations sought a confidence level of 0.95. Because clustered sampling would be done, a design effect of 1.5 was applied in rounds 1 to 3 and of 2 in round 4. The resulting sample size was multiplied by 37, the number of districts in Tamil Nadu as of October 2020, for rounds 1 and 3. In round 2, the multiple was 36 because Chennai was not sampled. In round 4, the multiple was 38, as one of the districts was split into two by round 4. This implied state-wide sample size targets were 26,651 in rounds 1 and 3, 25,931 in round 2, and 32,664 in round 4.

### Sampling strategy

The study selected participants in each district in five steps. *First*, districts were divided into rural and urban strata. District-wise sample-size targets were allocated to rural and urban strata in proportion to strata population. *Second*, rural and urban strata were divided into geographic clusters, defined as a village and street segments in rural and urban strata, respectively. *Third*, strata-wise sample-size targets were converted into cluster sample-size targets assuming 30 persons were sampled per cluster. *Fourth*, random sampling was used to select the targeted sample-size of clusters from each strata in each round. *Fifth*, up to 30 were sampled from each cluster use a random starting point, systematic sampling of households, and the Kish *(4)* method to select one participant per household. (Additional details are in the Supplement).

### Data collection

Each participant was asked to complete a health questionnaire (including questions on prior infections and vaccination) and provide 5ml venous blood collected in EDTA vacutainers. Serum was analyzed for IgG antibodies to the SARS-CoV-2 spike protein using either the iFlash-SARS-CoV-2 IgG (Shenzhen YHLO Biotech; sensitivity of 95.9% and specificity of 95.7% per manufacturer) *(5)* or the Vitros anti-SARS-CoV-2 IgG CLIA kit (Ortho-Clinical Diagnostics; sensitivity of 90% and specificity of 100% per manufacturer) *(6)*. We obtained data on each reported COVID-19 case and death through May 2022 from the Government of Tamil Nadu and Covid19Bharat.org *(3)* and on the number of tests done through January 2022 from the Government of Tamil Nadu.

### Statistical analysis

All statistics are calculated separately for each round unless otherwise indicated.

#### Seropositivity

The proportion of positive CLIA tests by district is obtained by estimating a logit regression of test result on district indicators and reporting the inverse logit of the coefficient for each district indicator. Observations are weighted by the inverse of sampling probability for their age and gender groups; the sampling probability here and below is based on population counts from the 2011 Indian Census. We reweight to match the 2011 Census because the Kish method ensures even (rather than representative) sampling by gender and age. Clustered standard errors are calculated at the cluster level.

#### Seroprevalence

Seroprevalence by district is estimated in two steps. <u>First</u>, we calculate the weighted proportion of positive tests at the district level. (We explain an exception for Chennai in round 1 and Virudhunagar in round 3 in the Supplement.) All samples in a district were tested using the same type of CLIA kit. We estimate a logit regression of test results on district indicators and take the inverse logit of the coefficient for each jurisdiction indicator. Observations are weighted by the inverse of sampling probability for their age and gender groups. Clustered standard errors are calculated at the cluster level. <u>Second</u>, for each jurisdiction, we predict seroprevalence using the Rogan-Gladen formula *(7)*, test parameters for the kit used in each jurisdiction, and regression estimates of seropositive proportion by jurisdiction.

State-level seroprevalence is obtained by aggregating the seroprevalence across districts weighted by 2011 Census data on the relative populations of districts.

Seroprevalence by demographic group is estimated in three steps. *First*, we calculate the proportion of positive tests at the jurisdiction-by-demographic group level in that round using logit regressions of test results on jurisdiction-by-demographic group indicators. Demographic groups indicators are sex x age for 6 age bins. Standard errors are clustered at the cluster level. *Second*, we predict district-by-demographic group level seroprevalence using the Rogan-Gladen formula. *Third*, we compute the weighted average of seroprevalence at the demographic-group level using as weights the share of demographic-group population in each district using data from the 2011 Indian census.

Seroprevalence by vaccine status in each of rounds 2 to 4 (when vaccines were available) is estimated in the same manner we calculate seroprevalence by demographic group in a round, except we replace demographic group by vaccine status.

Seroprevalence by urban status is obtained in the same manner as seroprevalence by demographic group, with two changes. First, we use the urban status of a cluster in lieu of demographic status of an individual at each step. Second, observations in our regression are weighted by inverse of the sampling probability for their urban status.

The size of a population that was seropositive by the end of a round is obtained by multiplying our seroprevalence estimates for the population in that round by the size of that population (as reported in the 2011 Census).

#### Undercounting of infections

The degree of undercounting of infections in round 1 is estimated by dividing the estimated number of people that are seropositive in the Tamil Nadu population by the number of government-reported cases in that population as of 1 week before the median sampling date of that round (23 October 2020). We focus on round 1 because vaccinations started between after round 1 and some seropositivity in rounds 2 to 4 is due to vaccination, not infections. The lag accounts for the delay, both between infection and seropositive status and between infection and prevalence testing. We calculate the Pearson’s correlation coefficient between undercounting rate and testing rate (tests per million as of median date of testing) by district.

#### Waning antibodies

We estimate the decline of antibodies after infection and in the absence of vaccination using district-level observations and a linear regression of district-level seropositivity in round 2 on district-level seropositivity in round 1. We focus on round 1 because no participants were vaccinated before round 1, meaning all seropositivity is due to infection. Observations are weighted in proportion to the population of each district in the 2011 Census. To address the possibility that decay is masked by new infections or vaccinations, we estimate a second specification that includes as controls a measure of the percent of population infected between round 1 and round 2 and the fraction of respondents who self-report vaccination. The measure of infection, which we call the “adjusted cases rate”, is the number of new confirmed cases per capita between rounds 1 to 2, adjusted by the infection undercount rate in round 1 (seroprevalence rate in round 1 divided by cases per capita until round 1).

We estimate the decline of antibodies following two doses of vaccination in two steps. *First*, we restrict the sample to individuals from round 4 who had been vaccinated with their second dose at least at least 20 days prior to biosample collection. The 20-day delay is intended to omit the period of time during which antibodies are climbing post-vaccination. We do not consider individuals from round 2 because we do not have their date of vaccination and from round 3 because so few individuals were vaccinated by that date. *Second*, we estimate a linear regression with an indicator for whether a person was seropositive as the dependent variable and the number of years (i.e., number of days/365) since dose 2 as the independent variable.

Observations are weighted to match age and gender proportions in the 2011 Census. To obtain plausibly causal estimates, we use age as an instrumental variable (IV) for the number of days since vaccination. The logic for this instrument is that Tamil Nadu prioritized individuals for vaccination based on their age, with older age persons given greater priority; to validate this instrument, we create a binscatter of days since vaccination on age among individuals with only 1 dose of vaccine and confirm that days since vaccination rises with age. The drawback of this instrument is that it is possible that antibody decay is directly a function of age *(8)*; therefore, IV estimates should be taken with a grain of salt.

#### Attribution to infection or vaccination

We attribute the change in seropositivity from round *t* − 1 to round *t* to changes in the levels of infections and of vaccination using the formula:

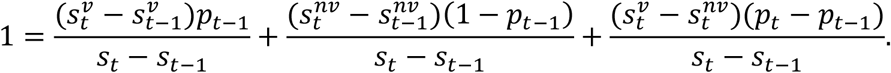

where *s*_*t*_ is the seropositivity rate in round *t, p*_*t*_ is the fraction of the sample vaccinated by round *t*, 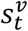 is the seropositivity rate among those vaccinated by round *t*, and 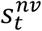 is the seropositivity rate among those not vaccinated by round *t*. The first term captures the share of the change in seropositivity *(s*_*t*_ − *s*_*t*−1_) attributable to changes in seropositivity rate among the previously vaccinated 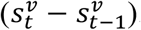. (This rate can change over time because antibodies levels may depend on the number of days since vaccination.) The second term captures the share attributable to infections among the previously unvaccinated (1 − *p*_*t*_). The third term captures the share attributable to changes in the vaccination rate. This captures both the effect of the increase in the vaccination rate *(p*_*t*_ − *p*_*t*−1_) and the change in seropositivity when one gets vaccinated 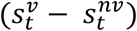. We calculate these components for the change in seropositivity from rounds 2 to 3 and from rounds 3 to 4.

Statistical tests comparing groups are performed using a two-sided Wald test with 95%. All statistical analyses were conducted with Microsoft Excel 365 (Microsoft, USA) and Stata 16 (StataCorp, USA). All plots were generated in R.

## Results

### Sample

In *round 1*, the study obtained results for 26,135 persons in 882 clusters (Table 1). The study could not sample 6 clusters and was unable to consent 324 persons in sampled clusters. One person aged 16 was incorrectly consented and dropped from the analysis. In *round 2*, the study obtained results for 21,992 persons in 746 clusters. (Chennai was not sampled.) The study could not sample 118 clusters and was unable to consent 388 persons in sampled clusters. Twenty-six persons age <18 were incorrectly consented and dropped from the analysis. In *round 3*, the study obtained results for 26,592 persons. The study could not consent 48 persons in sampled clusters. In *round 4*, the study obtained results for 32,244 persons. The study was unable to sample 13 clusters and could not consent 56 persons in sampled clusters. The final sample size per round was within the allowable 20% non-response rate.

**Table 1.**
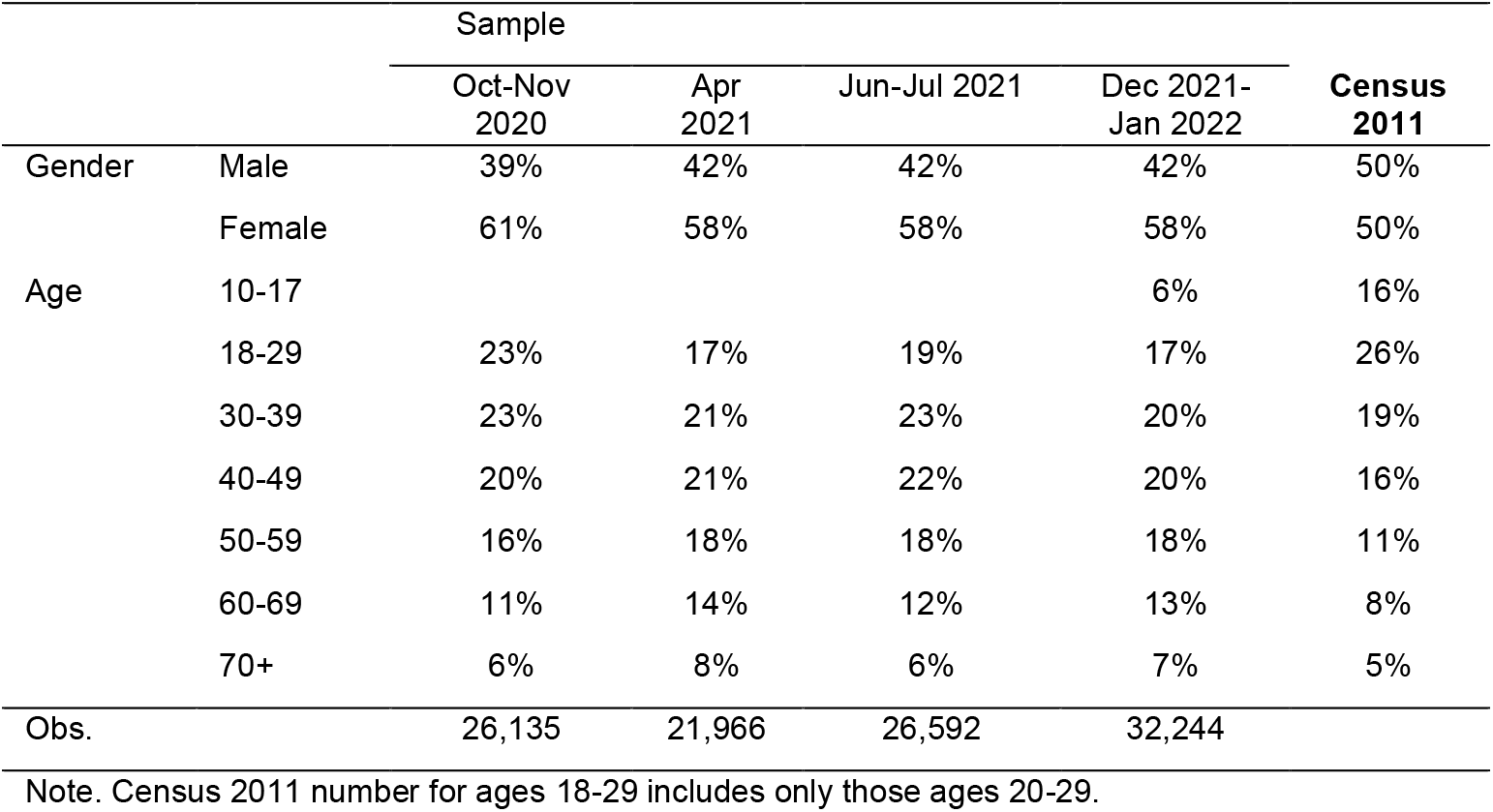
Demographics of sample, as compared to 2011 Census

|  |  | Sample |  |  |  | Census<br>2011 |
| --- | --- | --- | --- | --- | --- | --- |
|  |  | Oct-Nov<br>2020 | Apr<br>2021 | Jun-Jul 2021 | Dec 2021-Jan 2022 |  |
| Gender | Male | 39% | 42% | 42% | 42% | 50% |
|  | Female | 61% | 58% | 58% | 58% | 50% |
| Age | 10-17 |  |  |  | 6% | 16% |
|  | 18-29 | 23% | 17% | 19% | 17% | 26% |
|  | 30-39 | 23% | 21% | 23% | 20% | 19% |
|  | 40-49 | 20% | 21% | 22% | 20% | 16% |
|  | 50-59 | 16% | 18% | 18% | 18% | 11% |
|  | 60-69 | 11% | 14% | 12% | 13% | 8% |
|  | 70+ | 6% | 8% | 6% | 7% | 5% |
| Obs. |  | 26,135 | 21,966 | 26,592 | 32,244 |  |

Note. Census 2011 number for ages 18-29 includes only those ages 20-29.

Table 1 reports the demographic characteristics of the sample in each round. The sample has substantially more females and fewer persons aged 10-17 and 18-29 and more elderly persons than the general population.

### Seropositivity

State-level seropositivity was 33.0% (95% CI: 32.0-34.0%), 23.1% (95% CI: 22.2-24.0%), 67.5% (95% CI: 66.7-68.4%), and 88.3% (CI: 87.8-88.8%) in rounds 1, 2, 3 and 4, respectively (Table 2).

**Table 2.**
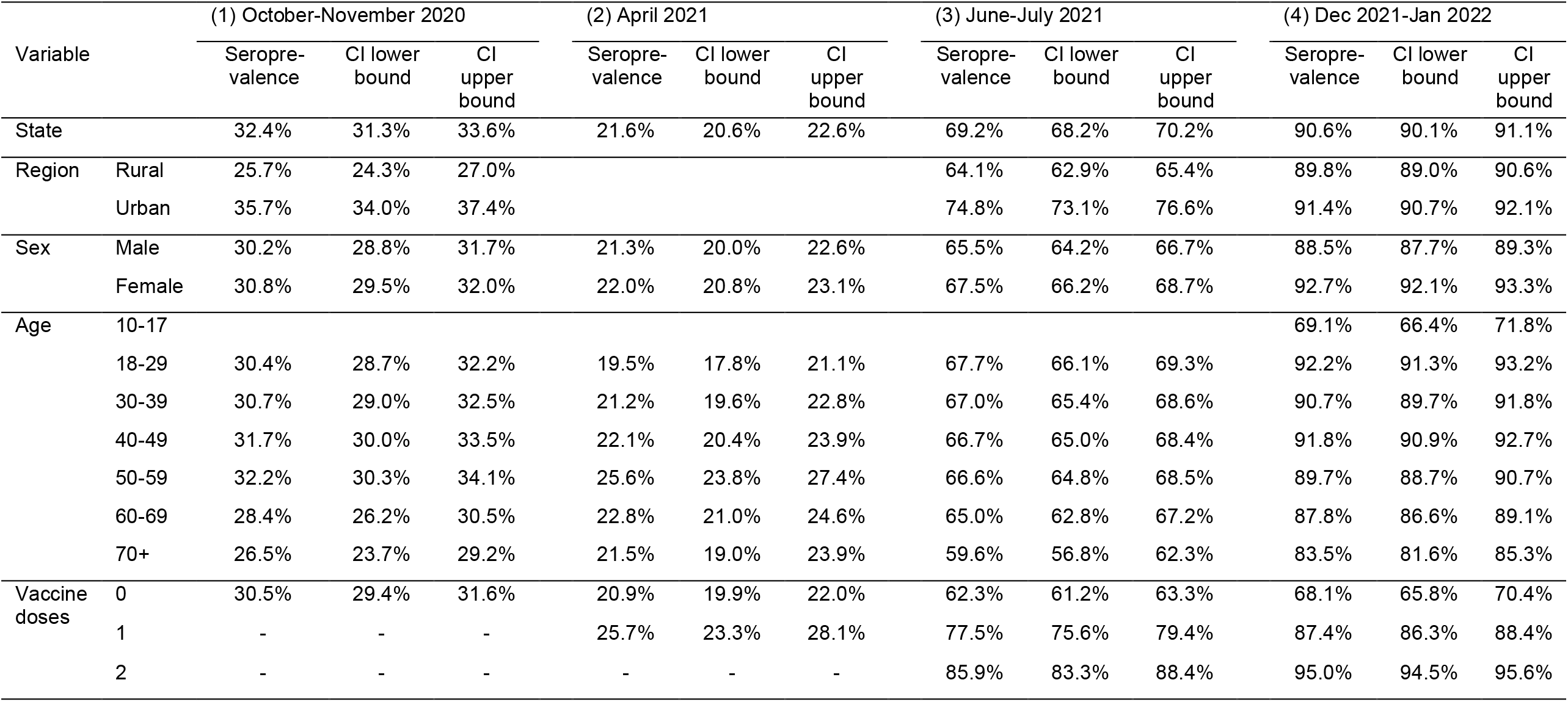
Seroprevalence by type of region, sex, and age

| Variable |  | (1) October-November 2020 |  |  | (2) April 2021 |  |  | (3) June-July 2021 |  |  | (4) Dec 2021-Jan 2022 |  |  |
| --- | --- | --- | --- | --- | --- | --- | --- | --- | --- | --- | --- | --- | --- |
|  |  | Seroprevalence | CI lower bound | CI upper bound | Seroprevalence | CI lower bound | CI upper bound | Seroprevalence | CI lower bound | CI upper bound | Seroprevalence | CI lower bound | CI upper bound |
| State |  | 32.4% | 31.3% | 33.6% | 21.6% | 20.6% | 22.6% | 69.2% | 68.2% | 70.2% | 90.6% | 90.1% | 91.1% |
| Region | Rural | 25.7% | 24.3% | 27.0% |  |  |  | 64.1% | 62.9% | 65.4% | 89.8% | 89.0% | 90.6% |
|  | Urban | 35.7% | 34.0% | 37.4% |  |  |  | 74.8% | 73.1% | 76.6% | 91.4% | 90.7% | 92.1% |
| Sex | Male | 30.2% | 28.8% | 31.7% | 21.3% | 20.0% | 22.6% | 65.5% | 64.2% | 66.7% | 88.5% | 87.7% | 89.3% |
|  | Female | 30.8% | 29.5% | 32.0% | 22.0% | 20.8% | 23.1% | 67.5% | 66.2% | 68.7% | 92.7% | 92.1% | 93.3% |
| Age | 10-17 |  |  |  |  |  |  |  |  |  | 69.1% | 66.4% | 71.8% |
|  | 18-29 | 30.4% | 28.7% | 32.2% | 19.5% | 17.8% | 21.1% | 67.7% | 66.1% | 69.3% | 92.2% | 91.3% | 93.2% |
|  | 30-39 | 30.7% | 29.0% | 32.5% | 21.2% | 19.6% | 22.8% | 67.0% | 65.4% | 68.6% | 90.7% | 89.7% | 91.8% |
|  | 40-49 | 31.7% | 30.0% | 33.5% | 22.1% | 20.4% | 23.9% | 66.7% | 65.0% | 68.4% | 91.8% | 90.9% | 92.7% |
|  | 50-59 | 32.2% | 30.3% | 34.1% | 25.6% | 23.8% | 27.4% | 66.6% | 64.8% | 68.5% | 89.7% | 88.7% | 90.7% |
|  | 60-69 | 28.4% | 26.2% | 30.5% | 22.8% | 21.0% | 24.6% | 65.0% | 62.8% | 67.2% | 87.8% | 86.6% | 89.1% |
|  | 70+ | 26.5% | 23.7% | 29.2% | 21.5% | 19.0% | 23.9% | 59.6% | 56.8% | 62.3% | 83.5% | 81.6% | 85.3% |
| Vaccine doses | 0 | 30.5% | 29.4% | 31.6% | 20.9% | 19.9% | 22.0% | 62.3% | 61.2% | 63.3% | 68.1% | 65.8% | 70.4% |
|  | 1 | - | - | - | 25.7% | 23.3% | 28.1% | 77.5% | 75.6% | 79.4% | 87.4% | 86.3% | 88.4% |
|  | 2 | - | - | - | - | - | - | 85.9% | 83.3% | 88.4% | 95.0% | 94.5% | 95.6% |

Seropositivity varied dramatically across districts in the first 3 rounds: from 11.9% (The Nilgris) to 49.7% (Perambalur) in round 1, 11.3% (Ramanathapuram) to 49.5% (Tiruvallur) in round 2, and 37.5% (Erode) to 81.9% (Chennai) in round 3 (Figure 2). Seropositivity converged by round 4, ranging from 82.9% (Tirupathur) to 94.3% (Thiruvarur).

**Figure 2.**
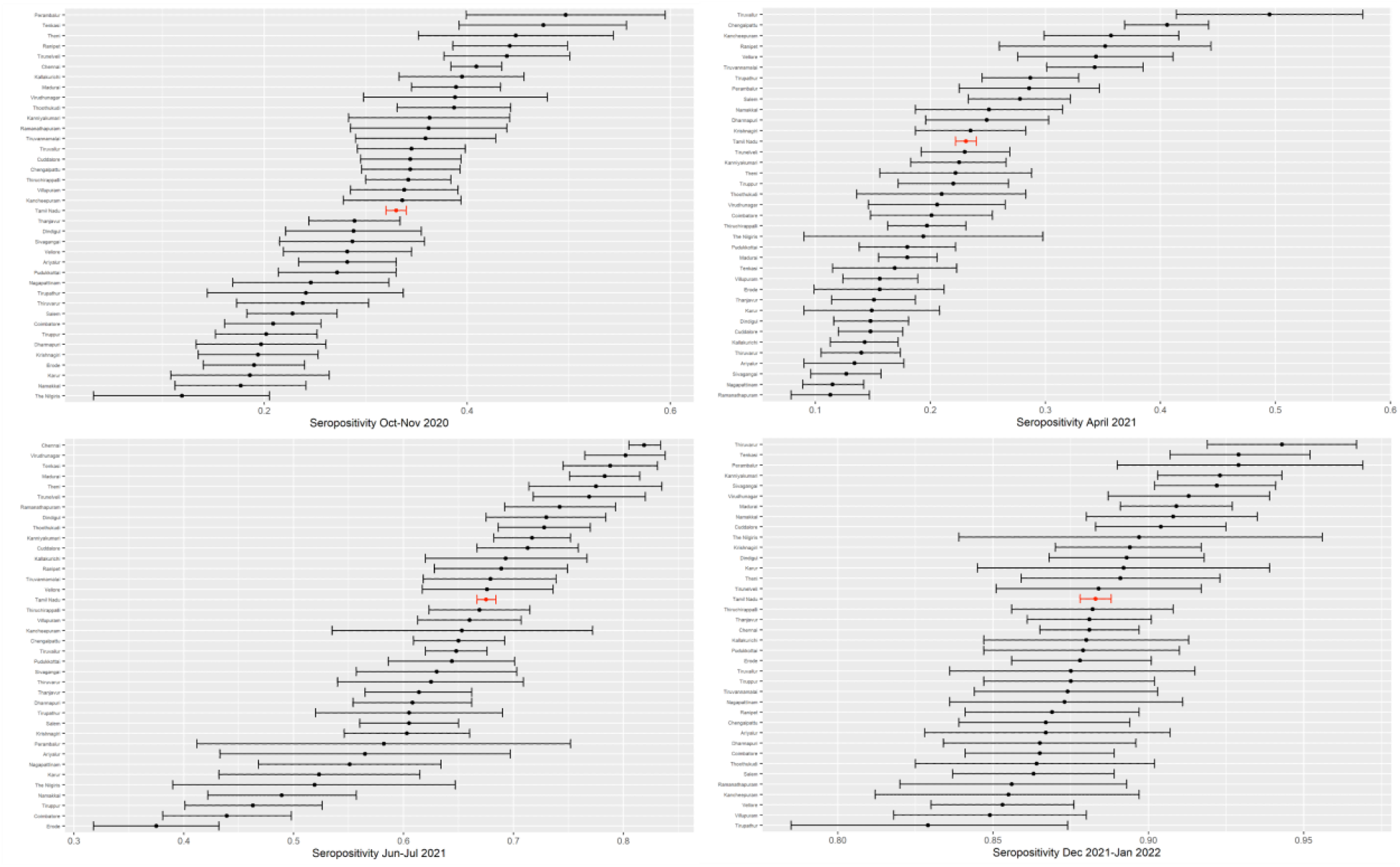
Proportion of positive CLIA tests by district

### Seroprevalence

State-level seroprevalence was 31.6% (95% CI: 30.3-32.7%), 22.9% (95% CI: 21.8-23.9%), 67.1% (95% CI: 65.9-68.3%), and 90.6% (CI: 90.1-91.1%) in rounds 1, 2, 3 and 4, respectively (Table 2). District-wise seroprevalence has a similar pattern to district-wise seropositivity (Supplement Figure 1).

Seroprevalence was significantly greater in urban areas than rural areas in rounds 1 (35.7% v. 25.7%, p<0.001) and round 3 (74.8% v. 64.1%, p<0.001) (Table 2). Urban classification of clusters was not available for round 2. By round 4, however, the gap has largely closed (91.4% v. 89.8%, p<0.001).

Seroprevalence is not substantially different across sexes (females v. males: 30.8% v. 30.2% in round 1; 22.0% v. 21.3% in round 2; 67.5% v. 65.5% in round 3; 92.7% v. 89.8%, round 4) (Table 2). While the round 4 difference is significant (p<0.001), it is still a small gap.

Seroprevalence is highest among older working-age populations in rounds 1 to 2 and among younger populations in rounds 3 to 4. Seroprevalence is significantly higher among older working-age populations than the elderly in rounds 1 to 3 (age 50-59 v. age 70+: 32.2% v. 26.5%, p=0.002 in round 1; 25.6% v. 21.5%, p=0.006 in round 2; 66.6% v. 59.6%, p<0.001 in round 3). Seroprevalence among young adult populations is significantly greater than among the elderly in rounds 3 to 4 (18-29 v. 70+: 67.7% v. 59.6%, p<0.001 in round 3; 92.2% v. 83.5%, p<0.001 in round 4) (Table 2). However, seroprevalence among the children aged 10-17 is lowest of all in round 4 (66.4%, p < 0.001 v. each other age group).

Seroprevalence is significantly greater among vaccinated populations (25.7% v. 20.9%, p<0.001; 80.0% v. 62.3%, p<0.001; and 93.1% v. 68.1%, p<0.001 in rounds 2, 3, and 4, respectively). Rounds 3 and 4 suggest that seroprevalence is increasing in number of doses taken (0 doses v. 1 dose: 62.3% v. 77.5% (p<0.001) in round 1, and 68.1% v. 87.4% (p<0.001) in round 2; 1 dose v. 2 doses: 77.5% v. 85.9% (p<0.001) in round 1 and 87.4% v. 95.0% (p<0.001) in round 2) (Table 2).

### Undercounting

The ratio of the number of infections implied by seroprevalence to confirmed cases ranges widely across districts, from 10 to 148 in round 1 (Supplement Table S 2). There is a significant negative correlation *(ρ*=-0.58, p<0.00) between COVID testing rate per thousand and the undercount rate in round 1 (Figure 3).

**Figure 3.**
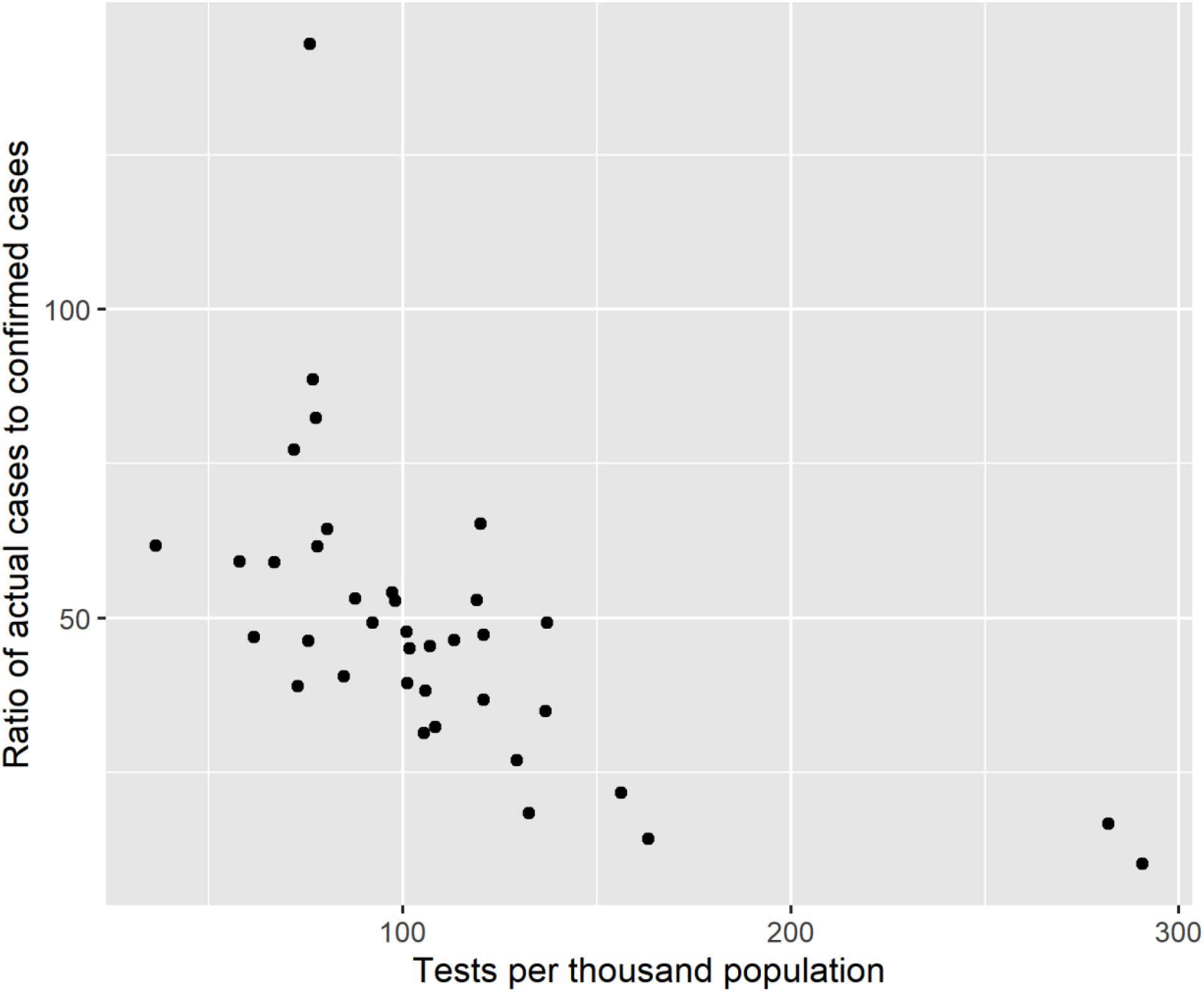
Relationship between rate of undercounting and testing rate in Round 1

### Waning antibodies

On average, district-wise seroprevalence rate in round 2 is 68.4% of the seroprevalence rate in round 1 in a district (Table 3), implying a 31.6% decline in seroprevalence, perhaps due to antibody waning. Across districts, the average adjusted cases rate is 9.35% between rounds 1 and 2 and on average 17.8% of sample members report being vaccinated with at least 1 dose by round 2. Adding (a) the district-level adjusted case rate to the regression to control for seropositivity due to new infection and (b) the self-reported vaccination rate to the regression to control for seropositivity due to vaccination yields a lower seroprevalence rate of 42.7% of round 1, implying a significantly larger 57.3% decline (p<0.001) in seroprevalence after infection.

**Table 3.** Decay of antibodies following natural infection

|  | (1) | (2) |
| --- | --- | --- |
|  | **Dependent variable:**<br>**Seroprevalence in round 2** |  |
| Seroprevalence in round 1 | 0.684***<br>(0.0635) | 0.427**<br>(0.129) |
| Adjusted case count |  | 1.204**<br>(0.370) |
| Fraction vaccinated<br>with 1 dose by round 2 |  | -0.143<br>(0.161) |
| N | 36 | 36 |

Notes. This table presents the results from regressing seroprevalence at the district level in round 2 against seroprevalence at the district level in round 1 and no constant. In column 2, the regression also includes the district-level adjusted case rate between round 1 and 2 to capture new COVID infections and the fraction vaccinated with 1 dose (self-reported) by round 2 to capture the new vaccination rate.

The annual rate of antibody decay after vaccination among individuals given 2 vaccine doses by round 4 is 16.3 percentage points after 1 year (Table 4). The seropositivity rate falls to zero within a year with the use of age as an instrumental variable for time since dose 2 to obtain causal estimates.

**Table 4.** Decay of antibodies following vaccine dose

|  | (1) | (2) |
| --- | --- | --- |
|  | **Dependent variable:**<br>**Seropositivity in round 4** |  |
| Years since dose 2 | -0.163***<br>(0.0125) | -1.001***<br>(0.150) |
| Constant | 0.960***<br>(0.00368) | 1.161***<br>(0.0359) |
| N | 15676 | 15676 |
| Total doses | 2 doses | 2 doses |
| Instruments | - | Age |

Notes. This table presents results from regressing seropositivity against years since second dose of vaccine. In the second column, years since last dose is instrumented by age.

### Attribution to infection or vaccination

Seropositivity increased by 42 percentage points (p.p.) between rounds 2, just before India’s second COVID wave, and round 3, after that wave (Table 5). Infections accounted for 74% of this increase. Increased seropositivity among those vaccinated by round 2 accounted for 23%, and new vaccinations accounted for just 2%. Seropositivity increased 23 p.p. from round 3-4, the period between India’s second and third wave. New vaccinations accounted for 65% of this increase. New infections and greater seropositivity among those vaccinated by round 3 accounted for 22% and 13% of this change.

**Table 5.** Attribution of seropositivity trends to infection or vaccination

| | Change in<br>seropositivity<br>$(s_t - s_{t-1})$ | Antibody response<br>given vaccination<br>$(s_t^v - s_{t-1}^v)p_{t-1}$ | Infection given no<br>vaccination<br>$(s_t^{nv} - s_{t-1}^{nv}) \times (1 - p_{t-1})$ | New vaccinations<br>$(s_t^v - s_t^{nv}) \times (p_t - p_{t-1})$ |
| --- | --- | --- | --- | --- |
| Round 2 to Round 3: |  |  |  |  |
| Percentage points | 42 | 9.6 | 31.1 | 1.1 |
| Percent of $(s_t - s_{t-1})$ | | 23% | 74% | 3% |
| Round 3 to Round 4: |  |  |  |  |
| Fraction | 23 | 3.0 | 5.2 | 15.1 |
| Percent of $(s_t - s_{t-1})$ | | 13% | 22% | 65% |

## Discussion

Serological surveillance suggests that officially confirmed cases dramatically underestimate the number of infections before vaccination. Statewide seroprevalence in round 1 implies that at least 22.6 million persons in Tamil Nadu were infected after Tamil Nadu’s first wave (by 30 November 2020). This estimate of actual infections is roughly 35 times larger than the number of confirmed cases by round 1 (674,802 cases by 16 October 2020) *(9)*.

Antibody decline after infection implies that even serological surveillance may underestimate actual infection rates before a vaccination campaign begins. Seropositivity declined between 31.6 and 57.3% over the roughly 6 months (170 days) between rounds 1 and 2. Evidence of cellular memory suggests that serological surveillance may also underestimate population immunity to COVID-19 *(10)*.

We also observe declining seropositivity – implying declining antibody counts -- after vaccination. However, we only observe individuals for at most 6 months after last dose. Therefore, one should not extrapolate from these data beyond one-half year. Moreover, participants with the greatest time since vaccination are also older, and older people may experience more rapid waning of antibodies *(8)*. Therefore, caution should be taken before extrapolating to younger populations.

Both infection and vaccination can contribute to seroprevalence. Between rounds 2 to 3, India experienced a second COVID-19 wave due to the Delta variant *(11)*. As a result, a majority of the increase in seropositivity from 23.1 to 67.5% was attributable to new infections. Between rounds 3 to 4, India ramped up its vaccination campaign and did not experience another COVID-19 wave. Therefore, most of the increase in seropositivity to 88.3% was attributable to vaccination.

The less-than-100% seropositivity rate or seroprevalence among the vaccinated, even conditional on days since vaccination (Supplement Figure S2), suggests that either some participants incorrectly reported being vaccinated or some doses may not have triggered a detectable antibody response.

Our study has several limitations. First, because antibody concentrations in infected persons decline over time *(12)*, our estimate of seroprevalence in round 1 may underestimate the level of prior infection and perhaps natural immunity.

Second, our estimate of antibody decline due to natural infection may be incorrect if our adjusted reported case rate does not accurately estimate the infection rate across districts. In that case, our control for infections in Table 3 is inadequate. The fact that a 1 percentage point increase in that adjusted rate is associated with a 1 percentage point higher seropositivity rate, however, suggests that the adjusted rate is a reasonable measure of infections.

Third, we may not accurately untangle seropositivity in round 3 that is due to infection versus due to vaccination. Our estimate of seropositivity among the vaccinated and among the unvaccinated during round 3 may be biased if there is selection into vaccination status that is correlated with seropositivity.

## Data Availability

These data are not publically available. 
However, researchers may approach the Government of Tamil Nadu to request access.  The decision to grant access will be made by the government.

## Author contributions

The study design and sampling were conceived, led and carried out by the Directorate of Public Health and Preventive Medicine, Government of Tamil Nadu, for the Government of Tamil Nadu’s Department of Health and Family Welfare. Data cleaning was performed by Vaidehi Tandel, Rajeshwari Parasa, and Sabareesh Ramachandran. Statistical analysis was conducted by Sabareesh Ramachandran and Anup Malani. Writing was done by Anup Malani, Sabareesh Ramachandran, and Stuti Sachdeva.

## Disclaimers

The opinions expressed by authors contributing to this journal do not necessarily reflect the opinions of the Centers for Disease Control and Prevention or the institutions with which the authors are affiliated.

## SUPPLEMENTAL MATERIALS

### Methods

#### Sample

Suspected or confirmed current or prior COVID-19 infection was not an exclusion criterion. If a participant was currently receiving medical care for COVID-19, a family member or proxy was used to complete the questionnaire on the participant’s behalf; however, the blood sample was taken from the participant.

#### Sampling strategy

The study selected participants in each district in five steps. *First*, districts were divided into rural and urban strata. All human settlements labeled villages in the 2011 Indian Census made up the rural strata. In rounds 1 to 3, the remaining settlements were the urban strata. (In round 4, the urban strata was further stratified into substrata comprised of municipal wards to make sure sampling was more geographically representative.) *Second*, rural and urban strata and substrata were divided into so-called clusters. In rural areas, each village was a single cluster. In urban areas, a street segment including between 50-500 households was called a cluster. *Third*, district-wise individual sample-size targets were converted into district-wise cluster sample-size targets assuming that 30 persons would be sampled per cluster. Clusters sample targets were assigned to rural and urban strata and substrata in proportion to the population of those strata. *Fourth*, simple random sampling was used to select the actual clusters to be sampled in accordance with cluster sample-size targets for each rural and urban strata or substrata.

*Finally*, within each cluster, a random GPS starting point was selected. One participant per household was sampled from households adjacent to that starting point until 30 persons consented within a cluster. Within each household, the participant asked to provide a biosample was selected via the Kish method *(4)*. If a participant refused, the survey went to next adjacent house until either 30 participants consented in the cluster or there were no more households in the cluster, whichever came first. The study asked participants using this process separately in each of the three rounds of survey; therefore, the participants sampled in each round may not be the same people sampled in other rounds.

#### Data collection

Blood was collected in EDTA vacutainers. Serum was isolated and stored in Eppendorf tubes. Serum was analyzed using either of two chemiluminescent immunoassay (CLIA) kits.

The first kit was the iFlash-SARS-CoV-2 IgG kit from Shenzhen YHLO Biotech. Per the manufacturer, it has a sensitivity of 95.9% (95% CI: 93.3-97.5%) and specificity of 95.7% (95% CI: 92.5-97.6%) *(5)*. Independent analysis estimated a sensitivity of 93% (95% CI: 84.3–97.7%) and specificity of 92.9% (95% CI: 85.3–97.4%) *(13)*.

The second kit was the Vitros anti-SARS-CoV-2 IgG CLIA from Ortho-Clinical Diagnostics. Per the manufacturer it has 90% sensitivity (95% CI: 76.3-97.2%) and 100% specificity (95% CI: 99.1–100.0%) *(6)*. FDA evaluation suggests it has 100% sensitivity (95% CI: 88.7-100%) and 100% specificity (95% CI: 95.4-100%) *(14)*. Independent analysis estimated that it has a sensitivity of 98.8% (95% CI: 92.9-100%) and specificity of 97.3% (95% CI: 85-100%) *(15)*.

All the samples in a district are analyzed using the same kit in a round, with the exception of Chennai in round 1 and Virudhunagar in round 3, where different HUDs used different kits. Table S 1 reports the test kit used in each district.

#### Statistical analysis

Generally, Nagapattinam district was split into Nagapattinam and Mayiladuthurai districts in March 2020, after the state started reported data on confirmed cases but before we conducted our serological survey. We aggregate these two districts together in our estimates of seropositivity and seroprevalence.

In Chennai, we do not have the population by HUDs. Since the samples were drawn proportional to population, we divide the district population across the HUDs in proportion to the sample size.

##### Seroprevalence

When estimating our district-level seroprevalence, the weights for our regression analysis employ data from the 2011 Census for the population in each age x gender category in each district. We estimate the sampling probability for demographic group (age category x sex) as the number of observations in that group in the sample in a district divided by the census population in that group in a district.

When estimating our urban- and rural-level seroprevalence, the weights for our regression analysis employ data from the 2011 Census for the population in each urban/rural category in each district. We estimate the sampling probability for urban/rural group as the number of observations in that group in the sample in a district divided by the census population in that group in a district.

We calculate the sampling probabilities for each regression observation at the level of 2011-defined districts (of which there are 32) rather than the 2020/21-defined districts (of which there are 37 or 38 depending on round), HUDs or clusters because the population is available only at the level of the old 32 districts. Likewise, we calculate district weights when we aggregate estimates across districts using the thirty-two 2001 districts. The 37 or 38, 2020/21 districts are all the same or bifurcations of the 2011 districts. Fortunately, in all bifurcated districts, the same kit was used. Therefore, we can combine all bifurcated districts into older 2011 districts for purposes of calculating sampling probabilities in regression analyses or weights when aggregating estimates.

Calculation of seroprevalence by district was modified in Chennai in round 1 and Virudhunagar in round 3. All samples in a district were tested using the same type of CLIA kit, except in Chennai and Virudhunagar, where all samples in a HUD were tested with the same type of kit. *The first step* was to calculate the weighted proportion of positive tests at the level of a health unit district (HUD), an administrative subset of districts. We estimate a weighted logit regression of test results on HUD indicators in Chennai and Virudhunagar and take the inverse logit of the coefficient for each jurisdiction indicator. Observations are weighted by the inverse of sampling probability for their age and gender groups. Clustered standard errors are calculated at the cluster level. *The second step*, for each jurisdiction, entailed predicting seroprevalence using the Rogan-Gladen formula *(7)*, test parameters for the kit used in each jurisdiction, and regression estimates of seropositive proportion by jurisdiction. In Chennai and Virudhunagar districts, we calculate seroprevalence at the district level as a weighted average of seroprevalence at the HUD level, using as weights the share of clusters in each HUD. We employ this approach to Chennai and Virudhunagar in estimators that use district-level seroprevalence.

##### Attribution of changes in seropositivity to infection or vaccination

The formula used to attribute the change in seropositivity rate across arms to infection and vaccination is derived as follows. First, we decompose the seropositivity rate *s*_*t*_ in round *t* into the fraction *p*_*t*_ of the sample vaccinated by round *t*, the seropositivity rate 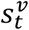 among those vaccinated by round *t*, and the seropositivity rate 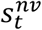 among those not vaccinated by round *t*:

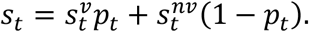

The difference between the seropositivity rate in round *t* and *t* − 1 is

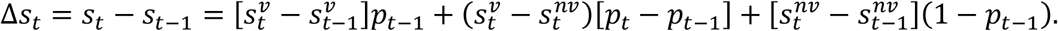

Dividing each side by the change in seropositivity across arms yields the fraction of changes in seropositivity attributable to changes in the seropositivity rate give vaccination, changes in the seropositivity rate given no vaccination, and, critically the increase in the vaccination rate:

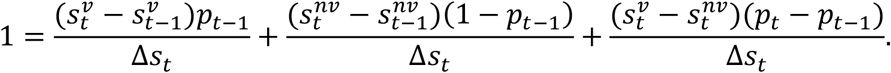

## Figures and tables

**Table S 1.**
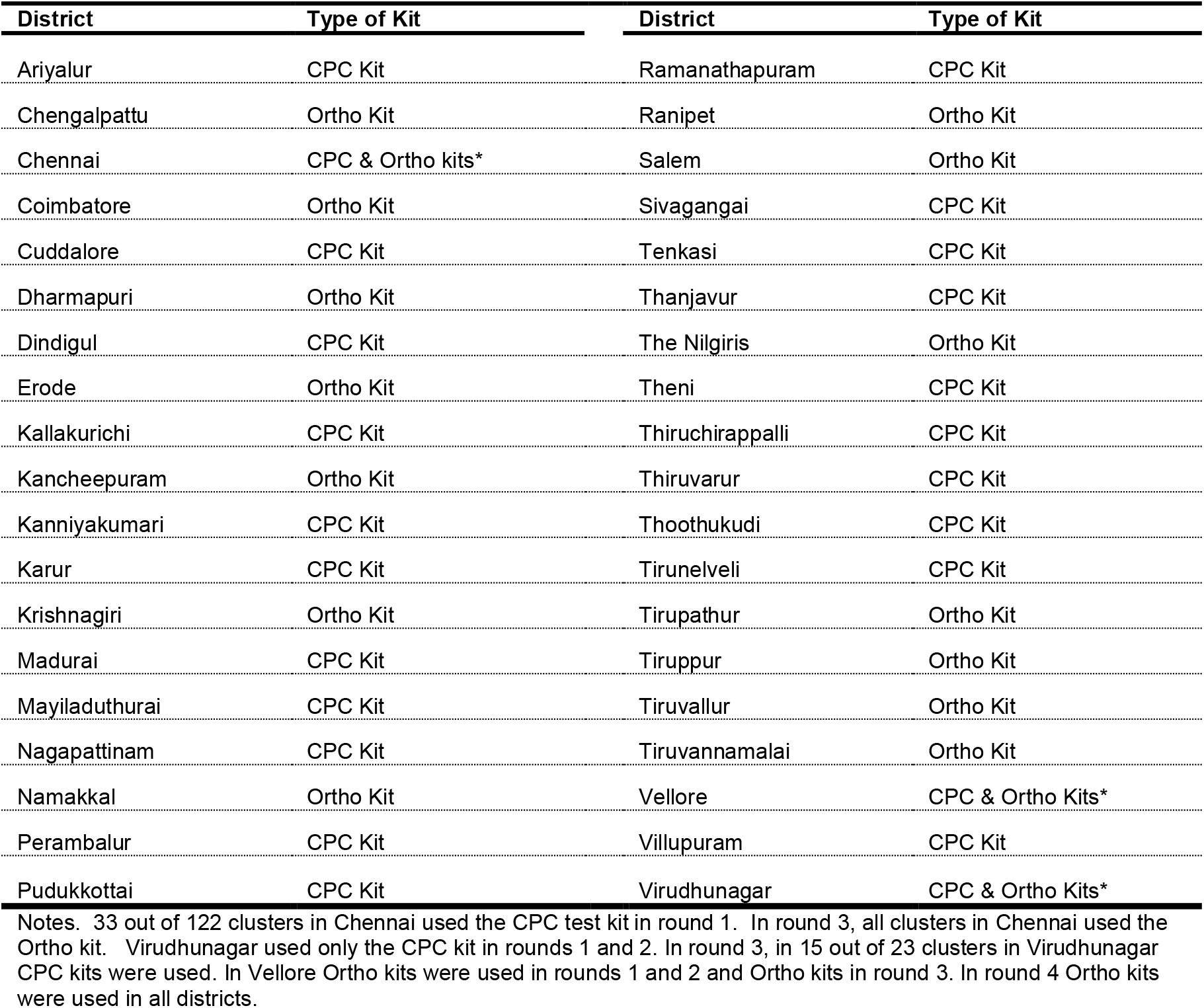
Test kit used in each district.

| District | Type of Kit | District | Type of Kit |
| --- | --- | --- | --- |
| Ariyalur | CPC Kit | Ramanathapuram | CPC Kit |
| Chengalpattu | Ortho Kit | Ranipet | Ortho Kit |
| Chennai | CPC & Ortho kits* | Salem | Ortho Kit |
| Coimbatore | Ortho Kit | Sivagangai | CPC Kit |
| Cuddalore | CPC Kit | Tenkasi | CPC Kit |
| Dharmapuri | Ortho Kit | Thanjavur | CPC Kit |
| Dindigul | CPC Kit | The Nilgiris | Ortho Kit |
| Erode | Ortho Kit | Theni | CPC Kit |
| Kallakurichi | CPC Kit | Thiruchirappalli | CPC Kit |
| Kancheepuram | Ortho Kit | Thiruvavarur | CPC Kit |
| Kanniyakumari | CPC Kit | Thoothukudi | CPC Kit |
| Karur | CPC Kit | Tirunelveli | CPC Kit |
| Krishnagiri | Ortho Kit | Tirupathur | Ortho Kit |
| Madurai | CPC Kit | Tiruppur | Ortho Kit |
| Mayiladuthurai | CPC Kit | Tiruvallur | Ortho Kit |
| Nagapattinam | CPC Kit | Tiruvannamalai | Ortho Kit |
| Namakkal | Ortho Kit | Vellore | CPC & Ortho Kits* |
| Perambalur | CPC Kit | Villupuram | CPC Kit |
| Pudukkottai | CPC Kit | Virudhunagar | CPC & Ortho Kits* |

Notes. 33 out of 122 clusters in Chennai used the CPC test kit in round 1. In round 3, all clusters in Chennai used the Ortho kit. Virudhunagar used only the CPC kit in rounds 1 and 2. In round 3, in 15 out of 23 clusters in Virudhunagar CPC kits were used. In Vellore Ortho kits were used in rounds 1 and 2 and Ortho kits in round 3. In round 4 Ortho kits were used in all districts.

**Table S 2.**
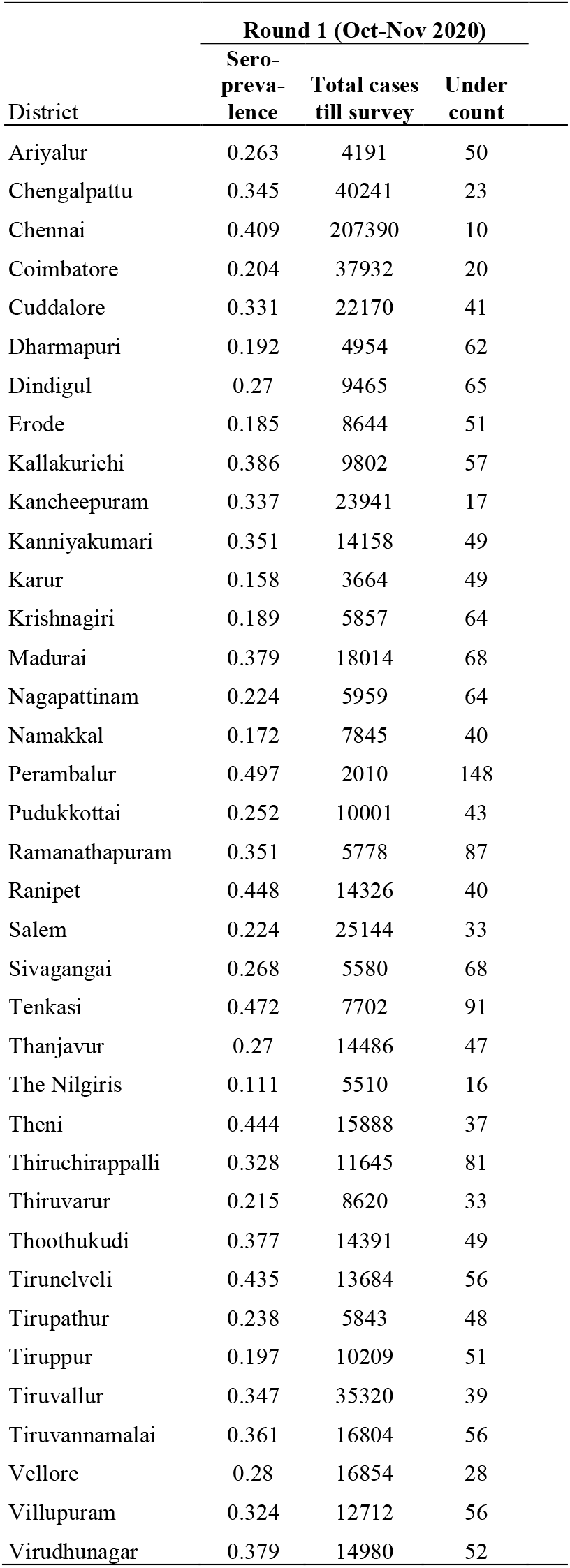
Confirmed cases and undercount of infections by district in round 1.

| District | Round 1 (Oct-Nov 2020) |  |  |
| --- | --- | --- | --- |
|  | Sero-prevalence | Total cases till survey | Under count |
| Ariyalur | 0.263 | 4191 | 50 |
| Chengalpattu | 0.345 | 40241 | 23 |
| Chennai | 0.409 | 207390 | 10 |
| Coimbatore | 0.204 | 37932 | 20 |
| Cuddalore | 0.331 | 22170 | 41 |
| Dharmapuri | 0.192 | 4954 | 62 |
| Dindigul | 0.27 | 9465 | 65 |
| Erode | 0.185 | 8644 | 51 |
| Kallakurichi | 0.386 | 9802 | 57 |
| Kancheepuram | 0.337 | 23941 | 17 |
| Kanniyakumari | 0.351 | 14158 | 49 |
| Karur | 0.158 | 3664 | 49 |
| Krishnagiri | 0.189 | 5857 | 64 |
| Madurai | 0.379 | 18014 | 68 |
| Nagapattinam | 0.224 | 5959 | 64 |
| Namakkal | 0.172 | 7845 | 40 |
| Perambalur | 0.497 | 2010 | 148 |
| Pudukkottai | 0.252 | 10001 | 43 |
| Ramanathapuram | 0.351 | 5778 | 87 |
| Ranipet | 0.448 | 14326 | 40 |
| Salem | 0.224 | 25144 | 33 |
| Sivagangai | 0.268 | 5580 | 68 |
| Tenkasi | 0.472 | 7702 | 91 |
| Thanjavur | 0.27 | 14486 | 47 |
| The Nilgiris | 0.111 | 5510 | 16 |
| Theni | 0.444 | 15888 | 37 |
| Thiruchirappalli | 0.328 | 11645 | 81 |
| Thiruvavarur | 0.215 | 8620 | 33 |
| Thoothukudi | 0.377 | 14391 | 49 |
| Tirunelveli | 0.435 | 13684 | 56 |
| Tirupathur | 0.238 | 5843 | 48 |
| Tiruppur | 0.197 | 10209 | 51 |
| Tiruvallur | 0.347 | 35320 | 39 |
| Tiruvannamalai | 0.361 | 16804 | 56 |
| Vellore | 0.28 | 16854 | 28 |
| Villupuram | 0.324 | 12712 | 56 |
| Virudhunagar | 0.379 | 14980 | 52 |

**Figure S1.**
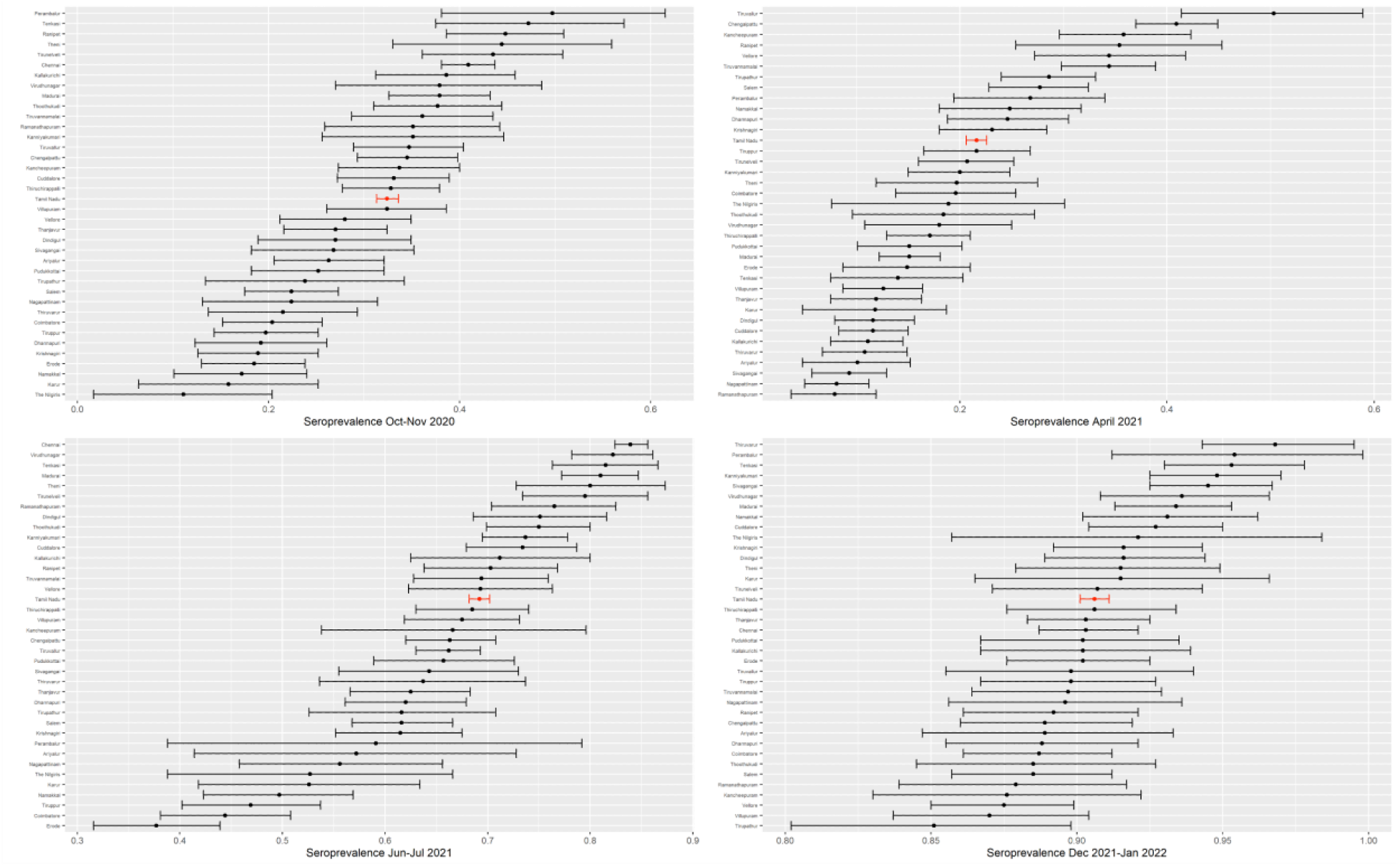
Seroprevalence by district and round

**Figure S2.**
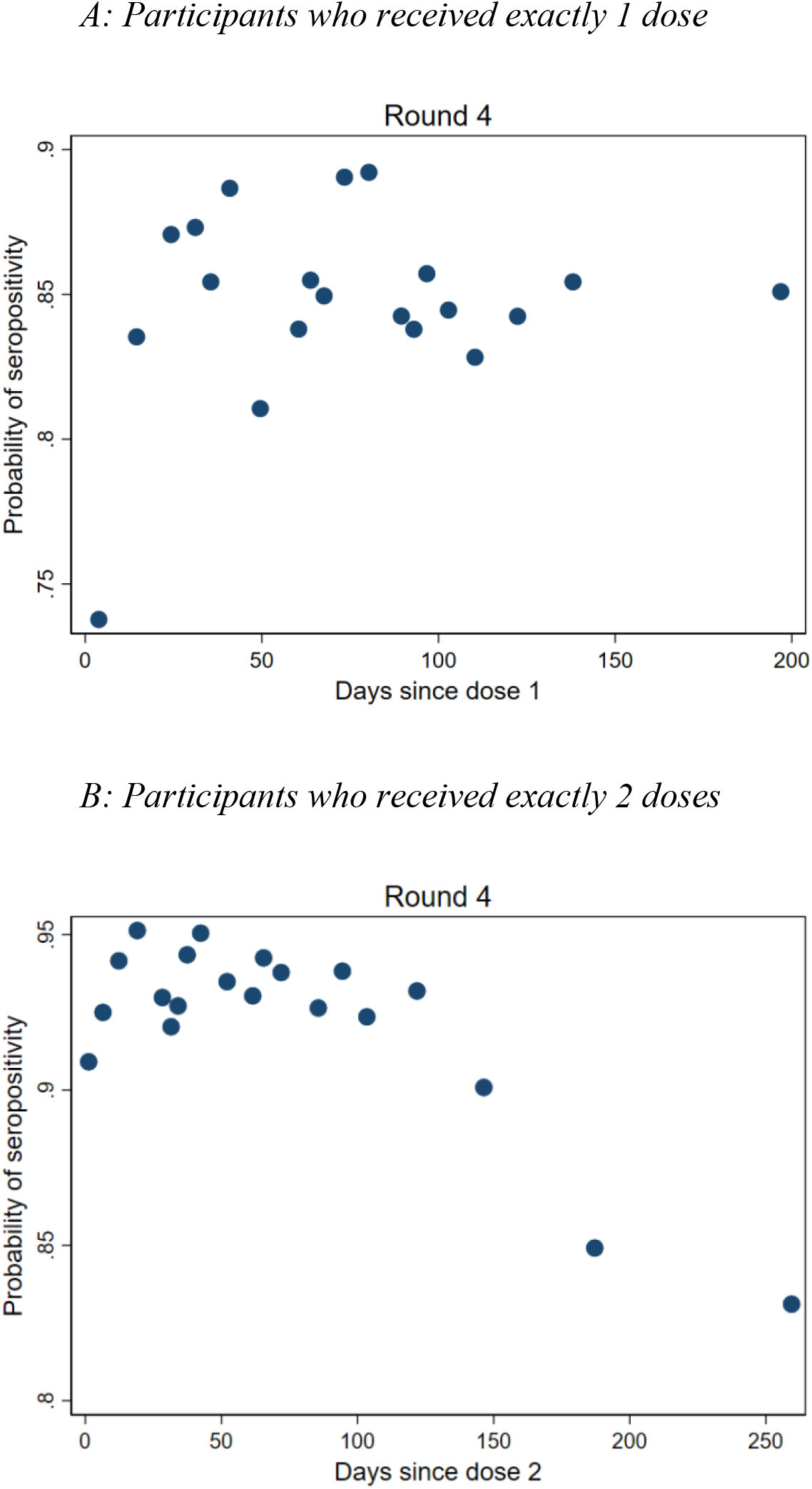
Decline in seropositivity by day since vaccine dose

## Notes

### Competing Interest Statement

The authors have declared no competing interest.

### Funding Statement

This study was funded by the Government of Tamil Nadu.

### Author Declarations

The Directorate of Public Health & Preventative Medicine, Government of Tamil Nadu, and the Institutional Ethics Committee of Madras Medical College, Chennai, India, gave ethical approval of this work.

### Summary of Updates

Data from 1 more round (4th round) of sero-surveillance in December 2021-January 2022 was added. In addition, we added a decomposition to examine the role of infection versus vaccination in driving changes in seropositivity between rounds.

